# Neuroimaging-based analysis of DBS outcome in pediatric dystonia: Insights from the GEPESTIM registry

**DOI:** 10.1101/2023.02.14.23285901

**Authors:** Bassam Al-Fatly, Sabina Giesler, Simon Oxenford, Ningfei Li, Johannes Achtzehn, Patricia Krause, Veerle Visser-Vandewalle, Joachim K. Krauss, Joachim Runge, Vera Tadic, Tobias Bäumer, Alfons Schnitzler, Jan Vesper, Jochen Wirths, Lars Timmermann, Andrea A. Kühn, Anne Koy, the GEPESTIM consortium

**Affiliations:** Charité-Universitätsmedizin Berlin, corporate member of Freie Universität Berlin and Humboldt-Universität zu Berlin, Department of Neurology, Berlin, Germany; Department of Pediatrics, Faculty of Medicine and University Hospital Cologne, University of Cologne, Cologne, Germany; Department of Neurosurgery, Hannover Medical School, Hannover, Germany; Department of Neurology, University Medical Center Schleswig Holstein, Lübeck Campus, Lübeck, Germany; Institute of System Motor Science, University Medical Center Schleswig Holstein, Lübeck Campus, Lübeck, Germany; Department of Functional Neurosurgery and Stereotaxy, Heinrich-Heine-University, Duesseldorf; Department of Stereotactic and Functional Neurosurgery, Faculty of Medicine and University Hospital Cologne, University of Cologne, Cologne, Germany; Department of Neurology, University Hospital of Marburg, Marburg, Germany

## Abstract

**Introduction:** Deep brain stimulation (DBS) is an established treatment in patients with pharmaco-resistant neurological disorders of different ages. Surgical targeting and postoperative programming of DBS depend on the spatial location of the stimulating electrodes in relation to the surrounding anatomical structures and on electrode connectivity to a specific distributed pattern of brain networks. Such information is usually collected using group-level analysis which relies on the availability normative imaging-resources (atlases and connectomes). To this end, analyzing DBS data of children with debilitating neurological disorders like dystonia would make benefit from such resources, especially given the developmental differences between adults and children neuroimaging data. We assembled pediatric, normative neuroimaging-resources from open-access neuroimaging datasets and illustrated their utility on a cohort of children with dystonia treated with pallidal DBS. We aimed to derive a local pallidal sweetspot and explore a connectivity fingerprint associated with pallidal stimulation to exemplify the utility of the assembled imaging resources.

**Methods:** A pediatric average brain template was implemented and used to localize DBS electrodes of twenty patients of the GEPESTIM registry cohort. Next, a pediatric subcortical atlas was also employed to highlight anatomical structures of interest. Local pallidal sweetspot was modeled and its degree of overlap with stimulation volumes was calculated as a correlate of individual clinical outcome. Additionally, a pediatric functional connectome of neurotypical subjects was built to allow network-based analyses and decipher a connectivity fingerprint responsible for clinical improvement in our cohort.

**Results:** We successfully implemented a pediatric neuroimaging dataset that will be made available to public use as a tool for DBS-analyses. Overlap of stimulation volumes with the identified DBS-sweetspot model correlated significantly with improvement on a local spatial level (R = 0.46, *permuted p* = 0.019). Functional connectivity fingerprint of DBS-outcome was determined as a network correlate of therapeutic pallidal stimulation in children with dystonia (R = 0.30, *permuted p* = 0.003).

**Conclusions:** Local sweetspot and distributed network models provide neuroanatomical substrates for DBS-associated clinical outcome in dystonia using pediatric neuroimaging surrogate data. The current implementation of pediatric neuroimaging dataset might help improving the practice of DBS-neuroimaging analyses in pediatric patients.

## Introduction

Image-based analyses of deep brain stimulation (DBS) effect has gained its popularity during the last decade^1,2^. In comparison to neuroimaging analyses in the adult populations, developmental differences could play a major role in analyzing pediatric neuroimaging data^3^. For instance, the basal ganglia and the thalamus widen and elongate with increasing age, in addition to many other morphological changes in gray and white matter^3^. Additionally, there is an uneven growth of brain structures that cannot be entirely represented with an adult average brain template^4,5^. In this regard, mapping DBS effects on a group-level necessitates a pipeline that takes a common pediatric template brain as a reference^6–9^, which has never been adopted in neuroimaging analyses of DBS in children^10–12^. Although transforming a pediatric brain image to an adult brain template is technically possible, the anatomical precision and visualization of basal ganglia structures may differ substantially, especially in regard to localizing DBS electrodes where millimeter differences matter^13,14^. Furthermore, the use of an adult template could lead to a higher degree of warping and distortion compared to a pediatric template^15,16^. On another aspect, drawing conclusions from DBS-associated remote networks has been shown to predict clinical improvement in a multitude of studies in adult populations^8,9,17–19^. Translating the concept of connectomic-DBS would then benefit from a pediatric connectome representative of age-respective developmental variances. An example of these variances is increased motor network functional connectivity (particularly in the basal ganglia) during childhood which correlates with increasing child-age^20,21^.

Neurological disorders can affect different age groups. Dystonia is a good example which can be manifested in different stages of the lifespan. Dystonia is characterized by sustained or intermittent muscle contractions causing abnormal, often repetitive, movements, postures, or both in one or more body regions^22^. On clinical grounds, dystonia is defined as isolated dystonia if it is the only feature with or without tremor, or as combined dystonia when associated with other movement disorders. The etiology of dystonia can be inherited, idiopathic, or acquired. Moreover, the pathophysiological background is complex in this network disease and involves abnormal inhibition, plasticity and other features on different levels of the nervous system^23,24^. Pediatric dystonia is a difficult condition with negative impact on patient’s quality of life and on caregivers^25,26^. Pharmacotherapy is very limited due to lack of effects or intolerable side-effects^27^. DBS has been established as a safe and effective treatment alternative for pharmacorefractory dystonia^28^. Apart from the adult population, multiple studies aimed at investigating the beneficial outcomes of DBS in children with idiopathic or inherited dystonia, with the globus pallidus internus (GPi) as the most common target for electrode implantation^29–31^. Despite several studies on optimal targeting for best clinical outcome in adults, there is very limited data investigating how neuroimaging-based analyses could improve targeting of electrodes and clinical outcomes after pallidal DBS in pediatric cohorts^10,11,32^. Therefore, it is of paramount importance to investigate whether such analyses could open a window on the local distribution and remote connections of efficacious pallidal stimulation in children with dystonia.

In this study, we sought to build a neuroimaging resource for image-based analyses in pediatric DBS population. We wanted to demonstrate its utility to map clinical effects of pallidal DBS in a sample of a well characterized pediatric example-cohort from the German Registry on Pediatric DBS (GEPESTIM)^33^. In order to do that, we took advantage of publicly available pediatric neuroimaging resources instead of using conventional adult resources. Electrode localization was visualized in relation to a novel pediatric basal ganglia atlas. DBS clinical effects were then mapped in respect to a local ‘sweetspot’ and a whole brain ‘network-fingerprint’.

## Methods

### Pediatric Neuroimaging Dataset Assembly

In order to overcome the co-registration/normalization bias that can be introduced by warping pediatric images to an adult brain template, we incorporated an unbiased pediatric MNI template^3^ in Lead-DBS pipelines^13^. To ensure coverage of the full span of pediatric age, a template representative of the age group 4.5-18.5 years was chosen. To comply with the routine of spatially normalizing individual brain images into adult MNI space in Lead-DBS, the asymmetric version of the aforementioned age-range template was chosen. This template has a 1 × 1 × 1 mm resolution and represents an average of 324 enrolled children. In addition, the template is available in multispectral versions (T1, T2 and proton density (PD)). All of them have been included in order to take advantage of the multispectral option of spatial normalization routines.

Visualizing DBS electrodes in relation to the surrounding anatomical structures is highly important to clinicians and researchers. A pediatric DISTAL atlas was introduced as a new atlas in Lead-DBS warping specific structures from the adult DISTAL atlas^34^ relevant to the current work. First, the pediatric MNI space was co-registered and normalized to the default adult space used in Lead-DBS, namely the MNI152 NLIN 2009b. Manual refinement of the normalization step was carried-on on structures incorporated in the pediatric DISTAL atlas with WarpDrive^35^. Specifically, the globus pallidus internus (GPi), the globus pallidus externus (GPe), the subthalamic nucleus (STN) and the red nucleus were included in the new pediatric atlas as DBS target structures (GPi/GPe) and to further assure quality of alignment (using the subthalamic and red nuclei). In addition to facilitating electrode visualization, this atlas would allow us to precisely assess the relation of locally mapped DBS effect to anatomical structures of interest (in this case the GPi/GPe complex) using sweetspot analysis. The resulting inverse warp field was used to extract atlas structures in the pediatric MNI space.

As one of the neuroimaging-based analyses is to delineate the distributed functional network associated with beneficial DBS therapy, we sought to create a normative resting-state functional MRI (rs-fMRI) connectome. To do so, we downloaded and processed an rs-fMRI dataset from a sub-cohort of the Consortium for Reliability and Reproducibility (CoRR)^36^, namely nyu2 sub-cohort, containing neuroimaging data collected from neurotypical adult and pediatric subjects (http://fcon_1000.projects.nitrc.org/indi/CoRR/html/nyu_2.html)^36^. Specifically, only data from 107 subjects of age 6-18 years were included (an earlier version of the connectome is mentioned in^37^, for demographics and imaging protocols please see^36^, for scans parameters see suppl. table 1). The participants were informed by the investigators to rest during the whole scanning period while the eyes are opened. All participants underwent an exhaustive, psychometric tests to determine their neurotypical development. MRI scanning were performed in two different sessions during two different dates to conform with the aim of the original study (CoRR). However, for the purpose of the current connectome aggregation, we exclusively used data from session 1 since only few children have completed two sessions.

**Table 1:**
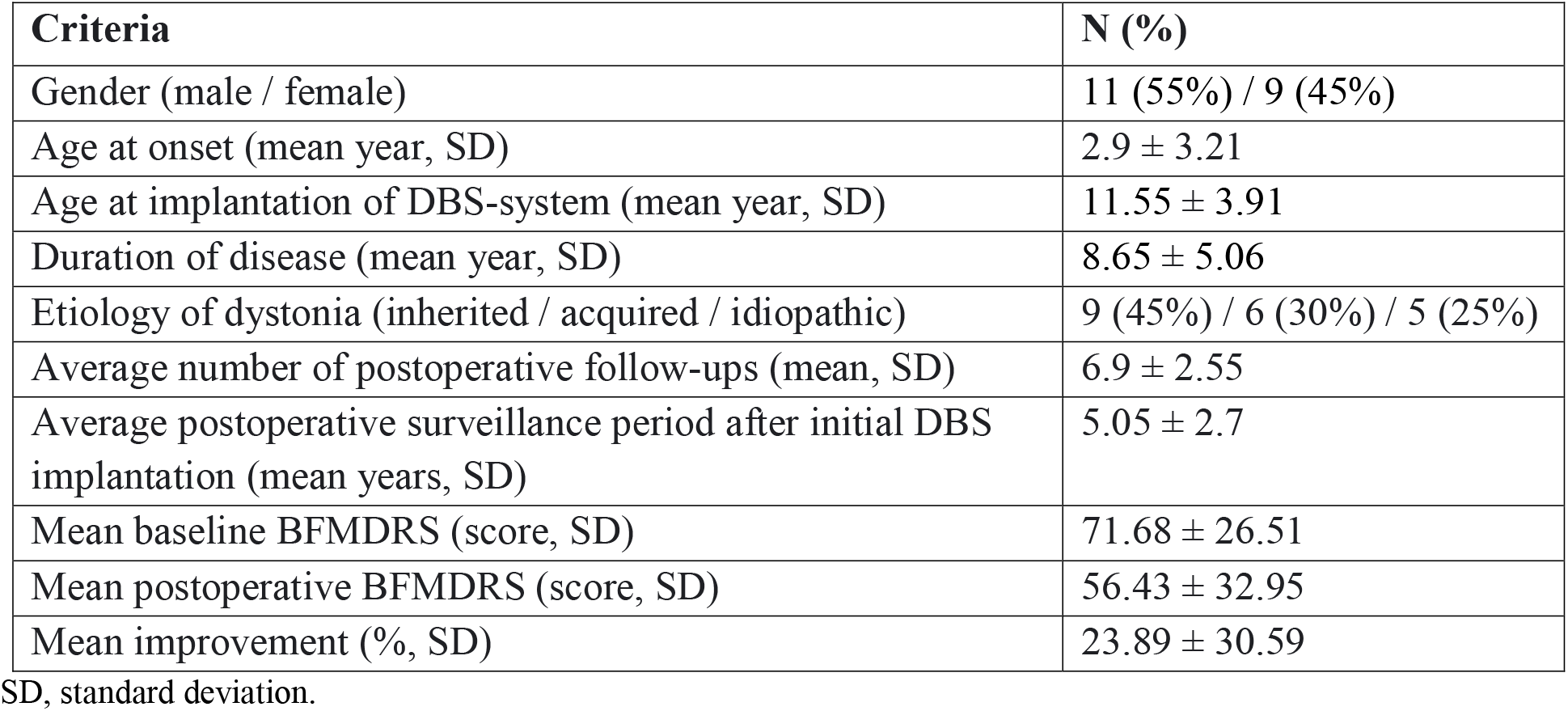
Patients demographic and clinical data.

| Criteria | N (%) |
| --- | --- |
| Gender (male / female) | 11 (55%) / 9 (45%) |
| Age at onset (mean year, SD) | $2.9 \pm 3.21$ |
| Age at implantation of DBS-system (mean year, SD) | $11.55 \pm 3.91$ |
| Duration of disease (mean year, SD) | $8.65 \pm 5.06$ |
| Etiology of dystonia (inherited / acquired / idiopathic) | 9 (45%) / 6 (30%) / 5 (25%) |
| Average number of postoperative follow-ups (mean, SD) | $6.9 \pm 2.55$ |
| Average postoperative surveillance period after initial DBS implantation (mean years, SD) | $5.05 \pm 2.7$ |
| Mean baseline BFMDRS (score, SD) | $71.68 \pm 26.51$ |
| Mean postoperative BFMDRS (score, SD) | $56.43 \pm 32.95$ |
| Mean improvement (% , SD) | $23.89 \pm 30.59$ |

SD, standard deviation.

### Normative connectome processing

Each subject’s anatomical and functional MRI data were first preprocessed using a collection of tools that belongs to different softwares (namely FSL, SPM and Lead-Connectome from Lead-DBS neuroimaging suite, https://www.lead-dbs.org/about/lead-connectome/). Slice time correction (FSL; https://fsl.fmrib.ox.ac.uk) was applied to the data. Realignment and initial motion correction of the resting-state functional magnetic resonance imaging (rs-fMRI) time series was then performed using *mcflirt* (FSL; https://fsl.fmrib.ox.ac.uk)^38^. Subjects were excluded if they have a framewise-displacement of > 0.5 mm in more than 50% of the volumes^39^ (seven subjects were excluded based on this criterion). Furthermore, detrimental motion effects were regressed out from the data using code implemented in *Lead-Connectome* (https://www.lead-dbs.org/about/lead-connectome/). Spatial smoothing was performed using a Gaussian kernel of 6 mm full width at half maximum, after which a high pass filter of 0.01Hz and a low pass filter of 0.08Hz was applied to the data in order to mitigate the effects of scanner drift and high frequency noise fluctuations,

Finally, we regressed out the average BOLD time series over cerebrospinal fluid and white matter^40^. To do this, the corresponding T1-weighted structural image for each subject was segmented using SPM “*newsegment*” function^41^. The resultant masks were linearly aligned (coregistered) to the rs-fMRI images, from which masks of white matter, cerebrospinal fluid and gray matter were obtained. The average signal over the cerebrospinal fluid and white matter masks was then calculated and regressed from the rs-fMRI time series via linear regression. Regression of global signal was also performed using Lead-Connectome Matlab code^42^. Normalization of functional volumes to the pediatric MNI space was then performed using FSL-FNIRT to nonlinearly warp the anatomical T1 images to the pediatric MNI space and later apply the warp to the coregistered rs-fMRI volumes. Following the normalization of each fMRI acquisition, a 285,903 × 180 matrix - containing the BOLD signal of every voxel (n = 285,903) for each volume in the time series (n = 180) - was computed using Lead-Connectome. The data were then masked to only include voxels within the brain in a readable format for seed-based connectivity analyses in *Lead-Mapper* (another toolbox from Lead-DBS neuroimaging suite).

### Study Cohort

Twenty children with a diagnosis of dystonia were selected from the GEPESTIM registry. Original trial data was provided by German DBS centers. After screening the available imaging data from the GEPESTIM cohort, the current data for analysis could be collected from five different neurological centers across Germany. Reasons for exclusion of data from the original GEPESTIM cohort were lack of pre- and/or post-operative imaging datasets or poor-quality scans (scans with artefacts) being insufficient for electrodes reconstruction and localization, lack of documented DBS settings corresponding to documented clinical scores, and insufficient period of postoperative clinical follow-up. Ethical approvals were provided by each participating center. Each patient was preoperatively assessed by an expert pediatric neurologist and at least 1 follow up was performed at 6 months or later postoperatively (given a latency of approximately 6 months for pallidal DBS to take effect). A preoperative and a postoperative Burke-Fahn-Marsden Dystonia Rating Scale (BFMDRS) score was collected for each patient^43^. DBS improvement was then calculated as the percentage ratio of the difference between the postoperative and the preoperative scores. Preoperative MRI scans were also acquired in each respective clinical center, in addition to a postoperative CT or MRI scan to confirm final DBS lead locations. There were regular postoperative follow-up visits to program the most clinically beneficial DBS settings. We used the most clinically stable DBS programming parameters. Detailed information and trial protocols can be found in the original GEPESTIM registry publication^33^.

### Electrode localization and stimulation volumes estimation

DBS electrodes were localized using the open-source software Lead-DBS (www.lead-dbs.org)^44^. The postoperative MRI or CT were co-registered to the corresponding preoperative MRI of each patient using Statistical Parametric Mapping software (SPM12; http://www.fil.ion.ucl.ac.uk/spm/software/spm12/)^45^ or the Advanced Normalization Tools (ANTs; http://stnava.github.io/ANTs/)^46^ respectively. The latter tools are integrated within Lead-DBS software. Next, all the co-registered patients’ images were warped to the pediatric MNI space using ANTs symmetric normalization (SyN) strategy. Both co-registered and normalized images were visualized and quality controlled for any mismatch. Importantly, we used a new feature of Lead-DBS WarpDrive^36^ to control for any minute difference in specific regions of the brain that still suffered malalignment to the pediatric MNI space by manually warping segments of the image. To minimize effects of brain-shift due to perioperative pneumocephalus, we applied brain-shift correction as implemented within Lead-DBS^13^. This strategy refines linear mappings between postoperative and preoperative scans using consecutive alignment routines focued on the target regions (basal ganglia). By doing so, nonlinear shifts introduced by pneumocephalus (usually present in frontal regions since patients are in supine position during scans) are substantially minimized. DBS electrodes were automatically pre-reconstructed using the PaCER algorithm^47^ for post-operative CTs and TRAC/CORE algorithm^13^ for post-operative MRI and later manually refined as implemented in Lead-DBS.

Stimulation volume surrounding active contacts was modeled using the SimBio/ FieldTrip approach^8^ implemented in Lead-DBS. Briefly, the electric fields (E-fields) were estimated in native space based on the individual optimized stimulation parameters using the finite element method. This was done by solving the static formulation of the Laplace equation on a discretized domain represented by a tetrahedral four-compartment mesh (composed of gray and white matter, metal and insulating electrode parts). Uniform conductivity of 0.14 S/m was applied to model gray and white matter. We adopted a simplified heuristic strategy, which thresholds electric fields at a vector magnitude above 0.2 V/mm^48^ and considers the resulting volume as “activated”. As a shorthand for the present manuscript, we refer to “stimulation volumes” as these thresholded volumes. Volumes were transferred to the pediatric MNI space using the deformation field calculated during spatial normalization.

### Sweetspot Analysis

Next, a group-analysis was performed on stimulation volumes to locally map DBS effect in the region of the pallidum. Previous studies used different sweetspot calculation methods^49^. However, in the current analysis, we opted to use a recently implemented method that was able to explain outcome in Parkinson’s patients implanted with DBS early in their disease process^6^. Briefly, every binarized-mask of each stimulation volume was weighted by its corresponding improvement score. Aggregated together, weighted volumes were then statistically tested with a one-sample t-test to test the average score of each voxel against zero. This allows to infer a sweetspot statistical map containing t-scores (T-model). To do so, voxels receiving contribution from less than 20% of total stimulation volumes were excluded to ensure robustness of the statistical test. We then calculated the sum of t-scores from the T-model voxels overlapping with each stimulation volumes. These sum values were then correlated with percentage improvement-scores using a 1000x permutation test (permuting improvement-scores). The permutation test was implemented to correct for multiple comparison and to test against null distribution and is free from assumptions about the distributions, which are typically violated in small sample sizes^50,51^. We repeated the same analysis on a subgroup of patients including cases with a diagnosis of inherited or idiopathic dystonia (n = 14). The latter analysis was meant to control for a possible effect of brain lesions in the acquired dystonia cases.

### Network Analysis

On the whole brain level, we wanted to test where the DBS electrodes should be connected to on a functional networks scale in order to obtain a beneficial DBS outcome. In a similar analysis to previous work^8,9^, we estimated functional connectivity profile of each stimulation volume to the rest of the brain using our assembled pediatric normative rs-fMRI connectome. In brief, average BOLD-signal of the bilateral stimulation volumes of each patient was correlated to each other voxel BOLD-signal in the brain of each of the connectome subjects. Average voxel-wise R values of 100 subjects were Fischer-z-scored to represent connectivity profile of each patient in our cohort. A voxel-wise correlation of connectivity strengths to corresponding DBS-related improvement values were then carried out across subject to yield a statistical R-map^8^. The R-map represents the optimal connectivity map which deciphers important regions of DBS functional network in children with dystonia. Spatial similarity of each stimulation volume associated connectivity profile to the R-map was then correlated with percentage improvement across all 20 subjects using a 1000x permutation test similar to the method above. Spatial similarities were calculated as the R coefficient of Pearson correlation between the voxel-wise connectivity values in the DBS-connectivity signature map and the voxel-wise R values in the R-map. The same analysis was repeated for the subgroup mentioned under sweetspot analysis section above (inherited/idiopathic group of n = 14).

### Data availability

Sensitive individual patients’ data cannot be shared for data protection reasons. All Lead-DBS codes can be accessed on https://github.com/netstim/leaddbs. Neuroimaging resources collected and processed in this manuscript including the pediatric MNI space/atlas and the connectome can be downloaded and queried from Lead-DBS interface.

## Results

### Patients’ characteristics

Patients included in the current analysis were part of a cohort of a registered trial (GEPESTIM)^33^. Five DBS centers from Germany provided data on 20 patients who underwent DBS until the age of 18 years between the years 2008-2020. In our cohort of 11 male and nine female subjects, the mean age of dystonia onset was 2.9 ± 3.21 years, the mean duration of disease at time of surgery was 8.65 ± 5.06 years and the mean age at DBS implantation was 11.55 ± 3.91 years. Average post-operative BFMDRS (56.43 ± 32.95 points) was significantly lower (*t*(19) = -2.99, *p* = 0.007, average percentage improvement = 23.89 ± 30.95 %) than pre-operative BFMDRS (71.68 ± 26.51 points). The average time to post-operative follow-up used to calculate percentage improvement in the study was 16.20 ± 12.89 months. Nine patients were diagnosed with inherited dystonia, six patients suffered from acquired dystonia, and in five patients no underlying cause could be identified (idiopathic dystonia). More precisely, the etiology of dystonia within the group of inherited dystonia was as follows: three patients were classified into the subgroup of DYT-TOR1A dystonia, DYT-KMT2B n = 1, DYT-SGCE n = 1, DYT-PRKRA n = 1, DYT-ANO3 n = 1, two patients showed mutations in the GNAO1 gene. Among the group of acquired dystonia, all patients experienced perinatal brain injury including perinatal asphyxia leading to dystonia. Overall, two patients with acquired and one patient with idiopathic dystonia were preterm (31, 32 and 36 weeks of gestation). In addition, we included one patient with hemi-dystonia, whereas 19 patients had generalized dystonia. Ten patients had isolated dystonia while the other ten presented with dystonia combined with another movement disorder. Pre- and postoperative BFMDRS scores were available for all 20 patients (see table 1 for demographics and clinical data, detailed individual patient related information can be found in suppl. table 2).

### Pediatric common brain space and subcortical atlas

An important step in the current study is the implementation of a common brain template of pediatric age to be used in DBS group-analysis. A multispectral version of the MNI pediatric space (Fig.1A) was incorporated and made openly distributed within the analysis pipeline of Lead-DBS software for future use in research. In addition, a pediatric atlas similar to the DISTAL adult atlas was created to highlight important subcortical structures surgically targeted or in the vicinity of the surgical target of DBS for dystonia. Precise transformation of the GPi, GPe, STN and red nucleus from the DISTAL adult to the current pediatric atlas was applied harnessing the manual refinement made possible by the WarpDrive tool (Fig.1B). This in turn ensured precision of visualization of the patients’ active contacts in relation to the anatomical structures in space (Fig. 1C).

**Figure 1:**
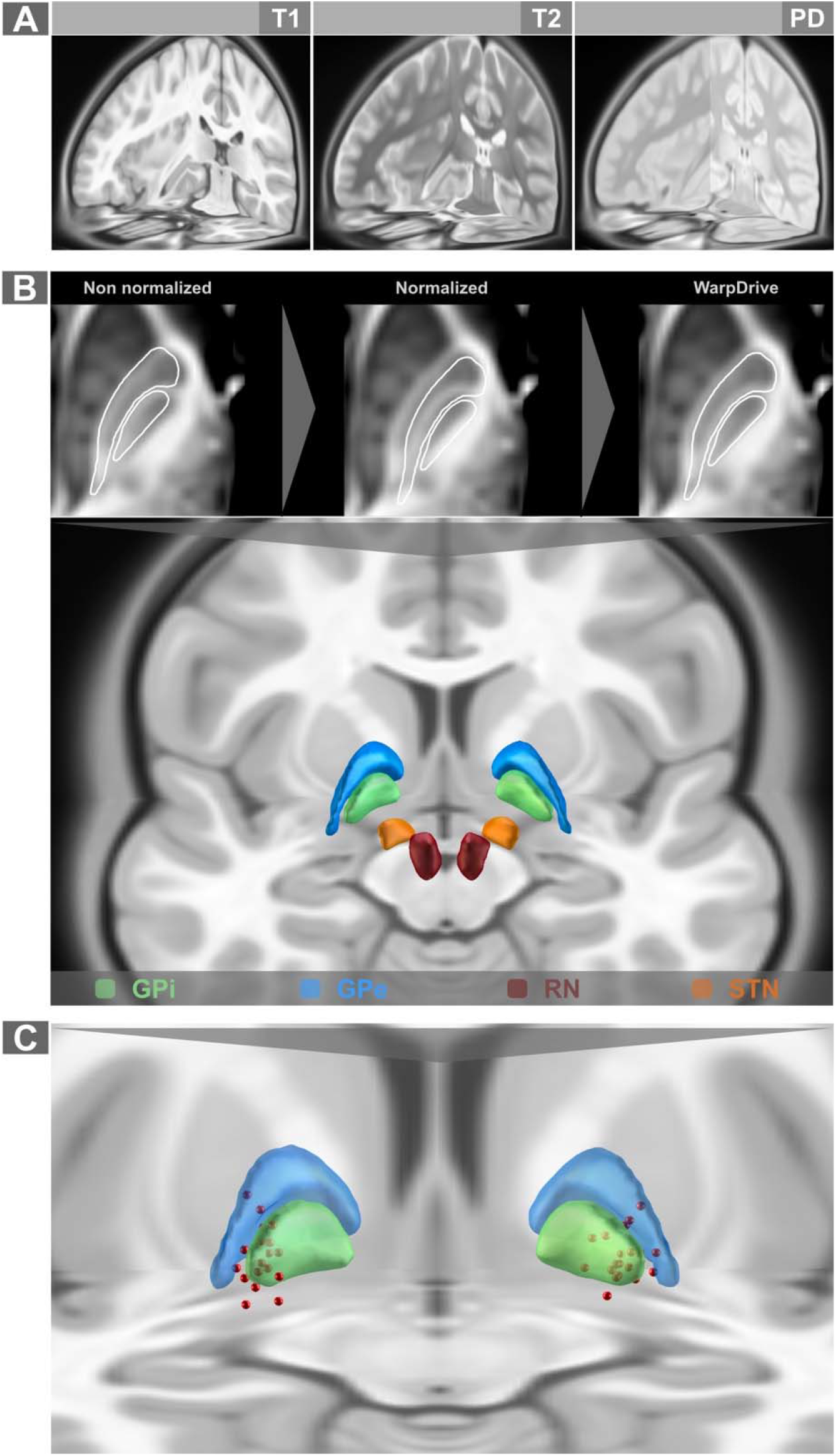
Pediatric standard (MNI) space and subcortical atlas. **A**. Multispectral MRI acquisitions of the pediatric MNI space^3^ were implemented as a standard space in Lead-DBS toolbox. **B**. Warping of relevant structures from the adult DISTAL atlas^34^ with fine-tuning using the WarpDrive tool was used to build the pediatric DISTAL atlas. **C**. Active contacts (red spheres) from the current GEPSTIM cohort included in the current study were demonstrated in relation to globus pallidus internus (green) and externus (blue). GPe, globus pallidus externus; GPi, globus pallidus internus; RN, red nucleus; STN, subthalamic nucleus.

### Degree of overlap with DBS sweetspot correlates with clinical improvement

The calculated statistical model (T-model) of the sweetspot was found to lie in the region of the GPi) with a tendency to encroach on ventral and lateral border of the GPi (on its interface with GPe) and slightly extending toward the subpallidal region (see Fig. 2A). Overlap of each combined bilateral stimulation volumes with the sweetspot correlated with corresponding DBS-associated clinical improvement (R = 0.46, *permuted p* = 0.019, Fig. 2B). The results were stable when repeating the analysis for inherited/idiopathic dystonia subgroup (R = 0.79, *permuted p* = 0.001, Suppl. Fig. 1).

**Figure 2:**
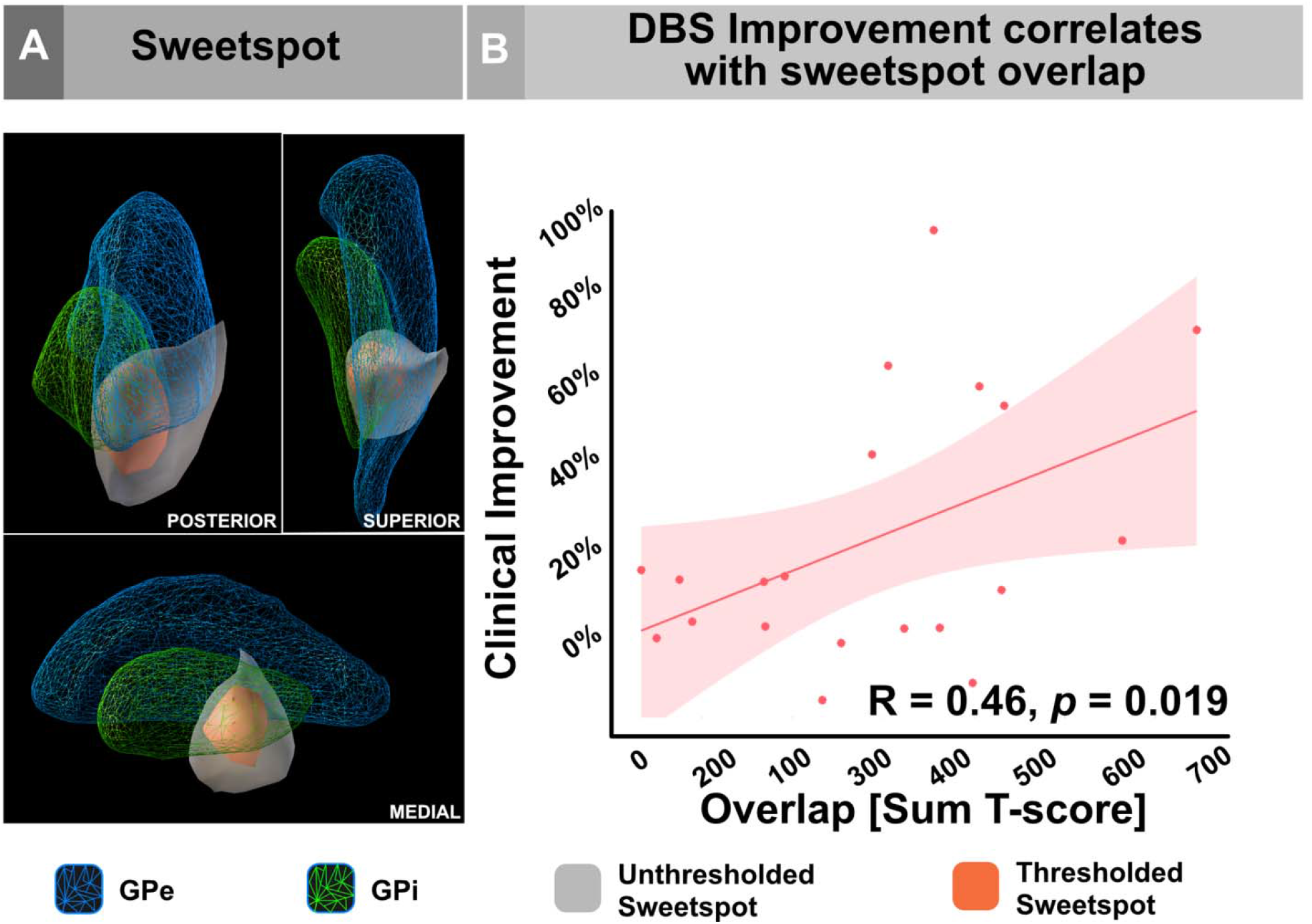
Sweetspot analysis. **A**. spatial location of the DBS sweetspot in relation to the right GPi/GPe complex from the current pediatric DISTAL atlas. Of note, the sweetspot spans a region lateral to the GPi on its interface to GPe in its posterior ventrolateral border. It also extends ventrally to subpallidal white matter. Sweetspot was presented as a t-score cluster (T-model, see methods) in unthresholded (red) and thresholded (grey, T ≥ 3) to demonstrate consistent extent regardless of threshold. **B**. Overlap of combined DBS stimulation volumes with the sweetspot (T-model) on local level correlated significantly with DBS-ON clinical improvement. The latter correlation was validated with permuting the percentage improvement scores 1000 times. GPe, globus pallidus externus; GPi, globus pallidus internus.

### A pediatric functional connectome

Rs-fMRI data from normally developing children were successfully processed and assembled as a ready-to use functional connectome for seed-based connectivity (Fig. 3). In order to pretest the robustness of the connectome, we seeded from a precuneal location (spherical seed of 4mm size). The seed was manually placed using Mango software (https://www.nitrc.org/projects/mango/) and later fed to Lead-Mapper to estimate the related-connectivity profile. The latter was in agreement with the canonical pattern of the default mode network^52^. This was a confirmatory step to ensure the vigor of the connectome. In addition, stimulation-related connectivity profiles were visually inspected to give an example on pallidal functional connectivity (see Fig 3B). It is worth mentioning, that the time-series data of this rs-fMRI connectome were sampled in the pediatric MNI space.

**Figure 3:**
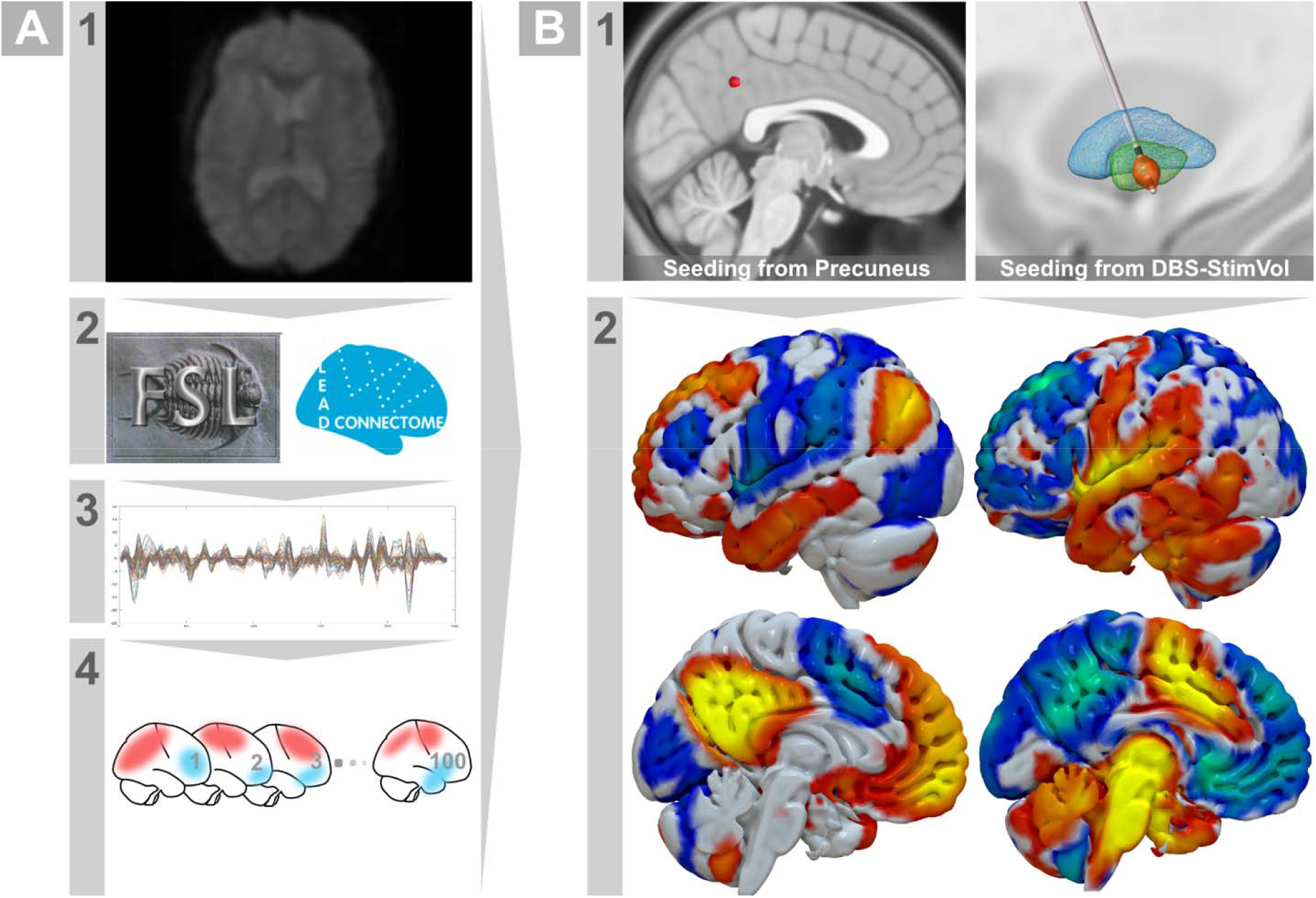
Resting-state normative pediatric connectome. **A**. Pipeline for connectome pre-processing. Raw fMRI scans (1) were downloaded and preprocessed with associated T1 MRI using FSL and Lead-Connectome tools (2) to extract BOLD signals (3) across 100 neurotypical children. Matrices of time-series were stored in a form usable by Lead-Mapper tool for seed connectivity analysis in each subject of the connectome (4). **B**. Estimation of average seed connectivity. (1) Example seed locations from the precuneus and a stimulation volume modeled from one of the patients in the current study cohort. (2) Using Lead-Mapper tool, estimation of seed connectivity profiles (averaged across 100 subjects in the connectome) successfully replicated the default mode network in case of the precuneal seed and relevant connectivity from GPi region was calculated using stimulation volume as a seed.

**Figure 4:**
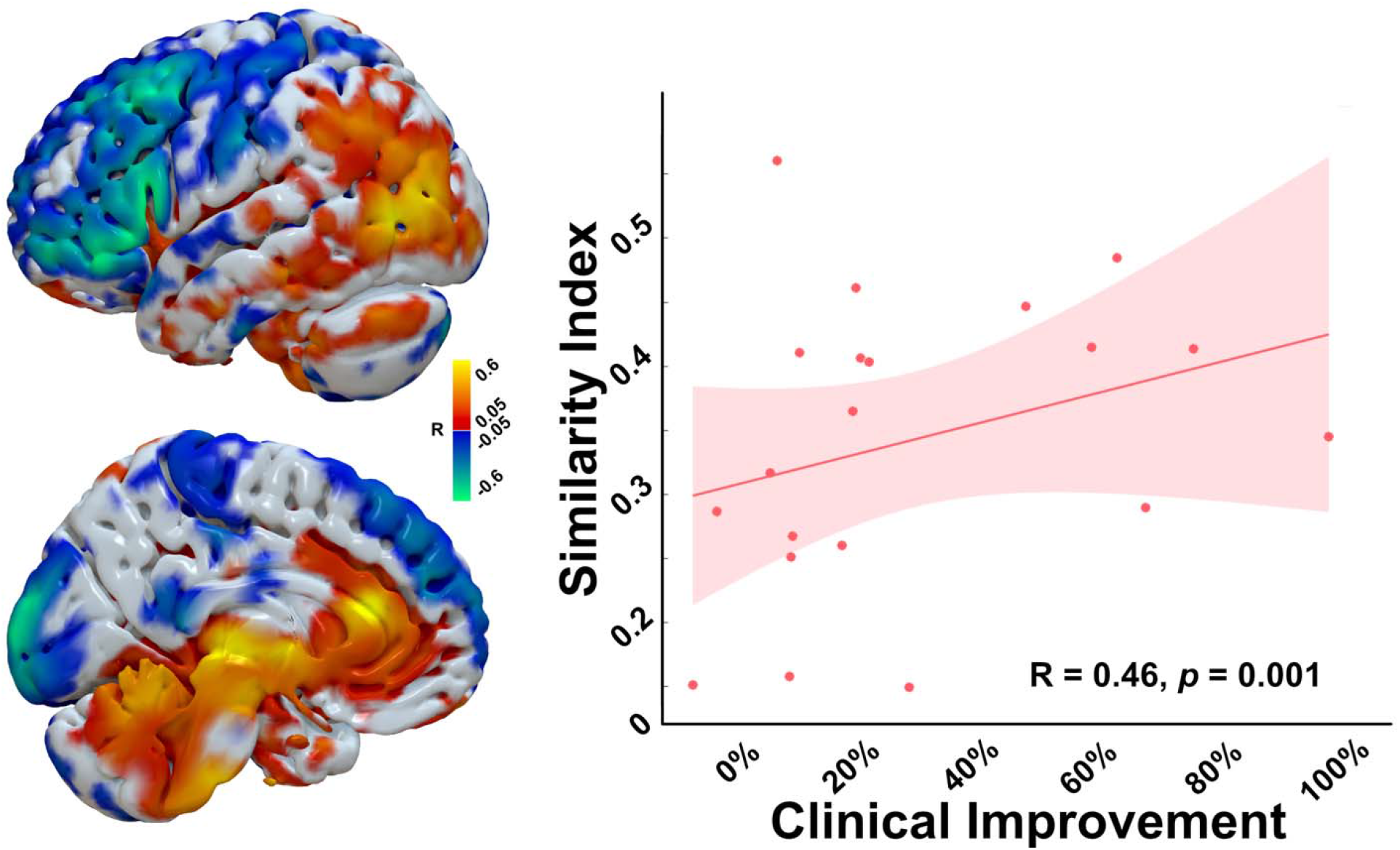
Functional connectivity fingerprint of beneficial pallidal DBS. R-map DBS-network model projected on a surface mesh of pediatric MNI space showing different regions and their relevance to DBS-outcome. Similarity of the DBS site associated whole-brain connectivity correlated significantly with clinical improvement across patients in our cohort.

### Connectivity pattern of DBS effect

In order to give further insight from a network perspective, a connectivity analysis was performed harnessing data from the pediatric functional connectome. While the sweetspot analysis looks at DBS effects from a localizationist point of view, functional network analysis extends on this concept and extracts related information from whole-brain regions that are indirectly and remotely (polysynaptically) connected to DBS electrode location. In this regard, the estimated group-level R-map demonstrated a peculiar topology highlighting connectivity to areas like the sensorimotor cortex, frontal cortex and posterior cerebellum to be negatively correlated with DBS effect (patients would worsen if their DBS electrodes connected more to these regions). On the other hand, connectivity to the parietal and anterior cingulate cortices were positively correlated with postoperative improvement. In addition, the brainstem and the medial and superior parts of the cerebellum were also similar pattern of connectivity correlation to imrpovement. In general, the similarity of the individual stimulation-related connectivity profile to the R-map model was significantly positively correlated with DBS improvement across our study cohort (R = 0.30, *permuted p* = 0.003). The model also yielded significant correlation using permutation testing when repeated on the inherited/idiopathic subgroup (R = 0.40, *permuted p* = 0.001, Suppl. Fig 2).

## Discussion

This study aimed at implementing a pediatric neuroimaging dataset and showcase the local and remote-network correlates of DBS therapeutic effects in children with dystonia that have been reported in a previous clinical trial (GEPESTIM)^53^. This allowed us to describe a sweetspot statistical-model representative of DBS effect by inferring a relationship between stimulation location and clinical improvement in our patients. In addition, a functional-network correlate of effective DBS therapy was elucidated. Taken together, the current findings shed light into how local and remote information can be exploited to explore DBS outcome in children using pediatric neuroimaging-analysis tools.

### Towards a refined neuroimaging approach in pediatric DBS

The current understanding of neurological diseases and their therapies depends to a high degree on group-analysis of data. When neuroimaging data are to be considered, a common brain template would be usually used to aggregate data and perform statistical inference^54^. To this end, the use of an age-specific brain template will give the opportunity to better estimate developmental differences across age-groups and account for any anatomical variance^54–56^. The use of the pediatric MNI template in the current study was a good example of the above concept. To our best knowledge, this is the first study to analyze DBS-related, pediatric neuroimaging data using such a pediatric imaging resource. Additionally, the use such a template helps in minimizing the amount of spatial deformation needed to warp subject images compared to adult templates^15,16,57^. The implementation of the pediatric MNI template has also lent the possibility to adjust an atlas of subcortical structures (namely the DISTAL atlas)^34^ from the adult MNI space to the pediatric one. The atlas contained the main DBS target featured by the GPi/GPe complex which is more relevant for a commonly targeted to treat childhood dystonia. Additionally, these and further subcortical structures (STN and red nucleus) worked as anchor points to refine spatial normalization using Lead-DBS toolbox^13^.

Given the current shift from location-to network-based understanding of neurological diseases and treatment^58^, the application of a normative pediatric functional connectome opens an avenue towards an age-respective connectomic approach in pediatric DBS studies. As structural and functional changes occur during development, connectivity patterns also change and reshape until the brain reaches maturity^20,21^. While previous work relied on adult connectomes analysis^10,11^, our pediatric connectome could pave the road to a future use of such a connectome in DBS and other pediatric research applications (for instance, mapping networks associated with specific lesions causing childhood neurological diseases)^59^. Furthermore, whole brain mapping of DBS therapeutic-effects allows for building a common network that can be targeted by invasive and non-invasive neuromodulatory techniques^60^. Taken together, the use of all three neuroimaging resources in the current work (a template, an atlas and a connectome) can be regarded as a further step towards a personalized neuroimaging analysis in pediatric-DBS research^61^.

### A pallidal DBS-sweetspot in children

Antidystonic pallidal sweetspots have been described previously in many adult DBS studies^17,62–65^. Two recent studies have reported efficacious stimulation site in dystonic children implanted in the GPi using different methodologies^10,32^. Of note, our sweetspot methodology has been recently used on an adult STN-DBS study^6^. Crucially, overlap of stimulation volume with the sweetspot “T-model” was significantly correlated with DBS-associated improvement across our cohort. This finding emphasizes the importance of treating the output of sweetspot methodology as a statistical model rather than a mere binary location^62^. The anatomical distribution of the sweetspot in the posteroventral GPi indicated a lateral location in the interface between GPi and GPe. This could explain the essence of the GPe and pallidal input fibers to GPi as an important role player in antidystonic DBS effect^66^. It is, however, difficult to hypothesize the implication of fiber tracts since our study did not use tractography as a method. Further experiments on how the effect of DBS can be relayed through fiber streamlines in the vicinity of the pallidum in children are needed given the possible developmental difference from adults^67^. That said, the proper use of normative structural connectome or atlas should be valuable, apart from well-acquired native diffusion-MRI of the patients. Another aspect is the ventral extension of the sweetspot to the subpallidal white matter. The latter could, to some degree, be regarded in line with the findings in a big adult cohort^62^, especially when compared to the results of generalized dystonia in Horn et al study^17^. Of note, previously published sweetspots were calculated using different statistical tests and were derived from different types of dystonia in adult populations.

### A functional connectivity correlate of antidystonic DBS-effect in pediatric patients

As has been already mentioned above, the use of network mapping of symptoms induced by brain lesions^58^, or in our case mapping dystonia improvement induced by DBS, has gained interest by scientists in recent years^68,69^. Apart from understanding the link between network modulation and disease pathophysiology on a distributed brain topology, network mapping allows for an efficient translation between different invasive and non-invasive therapeutic strategies by targeting common connectivity substrates as has been alluded to earlier^60^. Currently, there is a limited-applicability of non-invasive neuromodulation in children. This means, that the common network-target concept could be used in future trials on children. The current study highlighted a functional connectomic fingerprint which is partially in line with a relevant topology of previous work^17^. Specifically, the negatively correlated sensorimotor cortices were a central finding in multiple studies^17^. Furthermore, the somatosensory cortex has been repetitively shown to be of major influence on the mechanism of dystonia regardless of somatotopic distribution (focal or generalized)^70^. The current results demonstrate the role of cerebellar neuromodulation when mitigating dystonic symptoms with different therapies^71–73^. While the anterior cingulate cortex is not an obvious location associated with movement disorders at first sight, its contribution to motor control has been proved in primates and human behavior^74^. The connectomic fingerprint illustrated here overlaps partially with a recent one by Horn et al^17^ as mentioned above, using an adult functional connectome to map DBS therapeutic effect in an adult dystonic cohort. However, some differences in the topology could be due to the inherent characteristics of the study cohort (heterogeneous types of dystonia). Another explanation, could be that DBS-related connectivity pattern is age dependent. This, however, should remain highly speculative until a proper systematic analysis between adult and pediatric DBS cohorts using respective imaging-resources should be performed.

## Limitations

Firstly, small sample size of the dystonia cohort included in the current study is one of the limitations. However, dystonia is classified as a rare disease and its occurrence in childhood adds to the difficulties^75^. This aspect can be mitigated through collaboration with different clinical centers to overcome this particular aspect. Needless to say, we strived to use all possible data from our cohort to maximize number of participants included in the study. Secondly, the current cohort consisted of patients with different types of dystonia and different ages at surgery. These factors might have added to the heterogeneity of data, especially when taking into account that DBS effects do indeed depend on the underlying pathophysiology^76,77^. We accounted for the possible influence of acquired dystonia by repeating the sweetspot and network analyses while excluding them. Further bigger cohorts should allow for testing the latter assumption in a more systematic approach. Nevertheless, the current data should be seen as a foundation to the technical approach in pediatric DBS patients, although associated results should also be interpreted carefully. A third limitation is the implementation of a normative connectome for DBS-network mapping. Although the use of such connectomes has shown good performance in explaining network effect of DBS in different studies^8,9,17–19^, a possible comparison to patient-specific connectivity data is of importance^78^, especially that some patients have brain lesions that could impact the network as stated earlier. However, publicly available data are usually collected for research purposes and are of higher resolution than clinically collected data. Another specific limitation in children with movement disorders like dystonia is movement artefacts^79^. The latter could also favor the use of a more stable normative connectome. Additionally, average seed-based connectivity profiles derived from multiple subjects in a normative connectome can enhance the signal to noise ratio.

## Conclusions

We used a set of pediatric resources to perform neuroimaging analyses on a group-level in pediatric DBS patients. This enables us to identify a sweetspot of beneficial pallidal DBS effect, using a pediatric template and atlas, as well as an optimal whole-brain functional connectomic network, using a pediatric normative connectome. The latter corresponds to current knowledge about the pathophysiologic network model responsible for dystonia. Our findings confirm previous results and pave the road for future neuroimaging analyses in pediatric DBS research.

## Data Availability

Sensitive individual patients data cannot be shared for data protection reasons. All Lead-DBS codes can be accessed on https://github.com/netstim/leaddbs. Neuroimaging resources collected and processed in this manuscript including the pediatric MNI space/atlas and the connectome can be downloaded and queried from Lead-DBS interface.

## Acknowledgments

We would like to thank Dr Andreas Horn for his kind advice during the data analysis and pediatric resources assembly. We would also like to extend our thanks to the patients and their families. This work is funded by the Deutsche Forschungsgemeinschaft (DFG, German Research Foundation) – Project-ID 424778381 – TRR 295”. The GEPESTIM registry is funded by the Dr. Rita and Dr. Hans Günther Herfort Stiftung.

