## Supplementary material for "Neuroimaging-based analysis of DBS outcome in pediatric dystonia: Insights from the GEPESTIM registry": suppl. Fig. 1

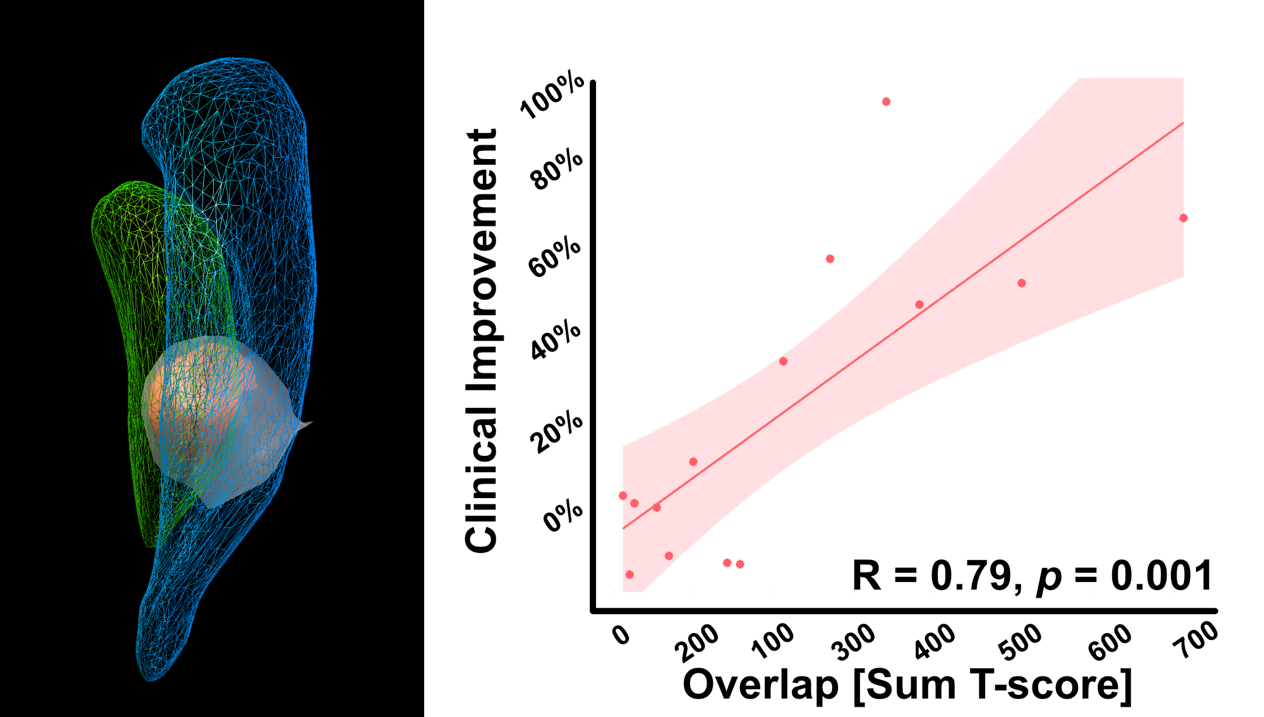

**Supplementary Figure 1**

Sweetspot overlap correlation for inherited and idiopathic cases. Left panel shows unthresholded sweetspot T-model in grey and thresholded (t > 2) in red together with a wireframe GPi (green) and GPe (blue) in pediatric MNI space.

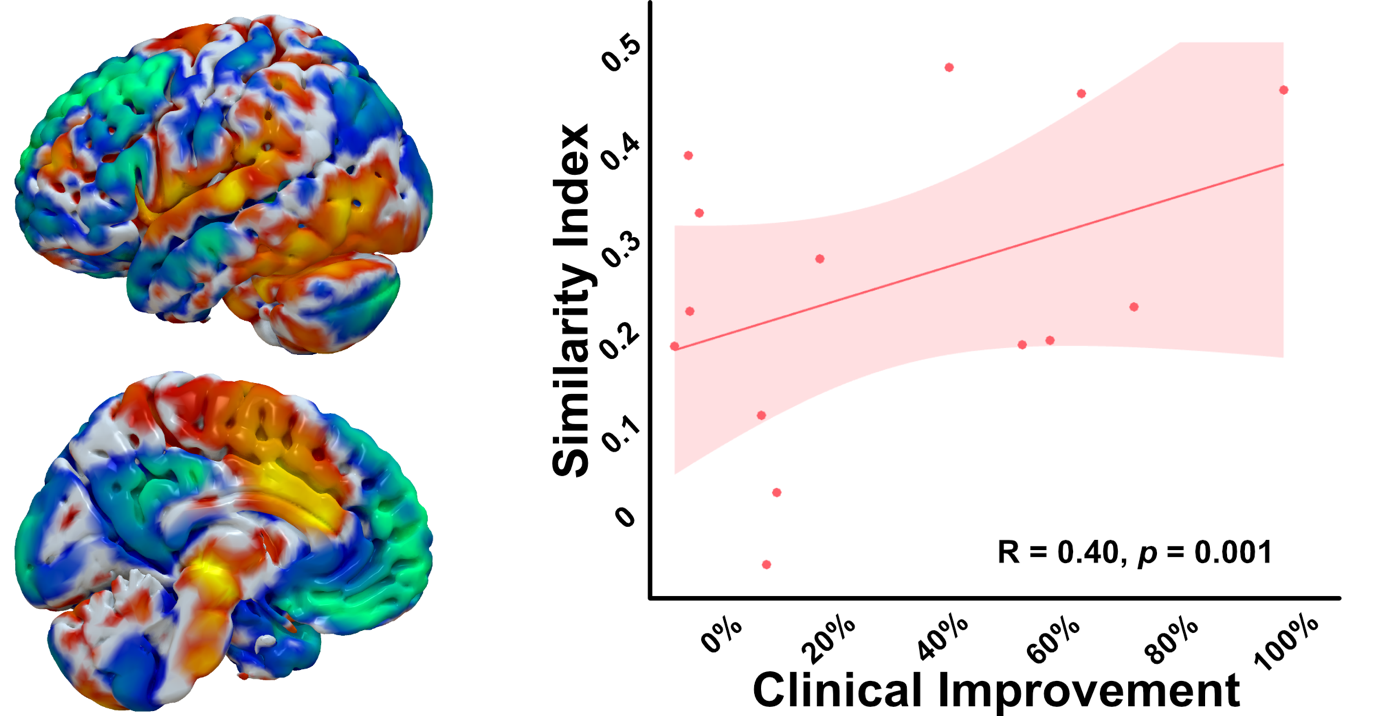

**Supplementary Figure 2**

Network correlation for inherited and idiopathic dystonia cases. Left panel shows the functional network correlate overlaid on a pediatric template surface model.
